# HIV Antiretroviral Therapy Scale-Up in Mozambique and Estimated Averted HIV Infections and Related Deaths, 2004–2023

**DOI:** 10.1101/2024.11.19.24317542

**Authors:** Sónia Chilundo, Irénio Gaspar, Ryan Keating, Joshua Fortmann, Dércio Filimão, Alexandre Nguimfack, Isabel Pereira, Charity Alfredo, Paula Samo Gudo, António Langa, Emilio Dirlikov, Seth Greenberg, Ishani Pathmanathan, Aleny Couto

**Author notes:** **Corresponding Author:** Sónia Chilundo.

## Abstract

**Introduction:** Mozambique is a HIV high-burden country. In 2004, Mozambique had an estimated HIV prevalence of 15.6% and 1.5 million people living with HIV (PLHIV). During same year the country started antiretroviral therapy (ART) scale-up supported by the U.S. President’s Emergency Plan for AIDS Relief (PEPFAR). We describe the ART scale-up and estimate averted HIV infections and HIV-related deaths.

**Methods:** We analyzed PEPFAR Monitoring, Evaluation, and Reporting program data from fiscal year 2004 to 2023. Before October 2018, PLHIV on ART were defined as clients with ≤90 days since last missed visit; in October 2018, this changed to ≤28 days. Viral load suppression (VLS) rate was calculated as PLHIV at PEPFAR-supported sites with VLS (<1,000 viral copies/mL) among those tested in previous 12 months. We used the 2024 United Nations Programme on HIV/AIDS (UNAIDS) Spectrum AIDS Impact Model and Goals Age Structured Model (ASM) to estimate HIV infections and HIV-related deaths averted.

**Results:** By September 2023, 2.1 million PLHIV in Mozambique were receiving PEPFAR-supported ART (1.4 million women [66%]; 1.9 million aged ≥15 years [95%]), a 650-fold increase from 3,226 in March 2004. The VLS rate increased from 62% (78,808/126,212) in 2017 to 94% (1,235,626/1,318,544) in 2023. During 2004–2023, ART scale-up helped avert an estimated 1.6 million HIV infections, including over 350,000 infections among HIV-exposed infants, and 1,070,295 HIV-related deaths.

**Conclusion:** The over 20 years of Mozambique’s scaled up ART effort has greatly expanded effective treatment and averted many new infections and HIV-related deaths. While progress has been impressive, gaps remain toward ending HIV as a public health threat.

## Introduction

In 2022, there were an estimated 39 million people living with HIV (PLHIV) worldwide, with 1.3 million new infections and 630,000 AIDS-related deaths.^1^ Despite global progress, HIV remains a leading cause of mortality in low-income countries, especially those in sub-Saharan Africa.^2^ Antiretroviral therapy (ART) reduces morbidity and mortality and prevents sexual transmission once a PLHIV’ viral load is suppressed to undetectable levels (<200 viral copies/mL).^3^ Globally, scale-up of ART reached almost 30 million PLHIV by 2022, and many countries are reaching the Joint United Nations Program on HIV and AIDS (UNAIDS) 95-95-95 targets for ≥95% of PLHIV to be diagnosed, ≥95% of those diagnosed to be on ART, and ≥95% of those on ART to be virally suppressed (<1,000 viral copies/mL).^1^

In Mozambique, HIV was first identified in 1986.^4^ Given resource constraints, Mozambique’s early response to HIV/AIDS focused on surveillance, prevention, and extremely limited access to treatment.^4^ In the early 2000s, the scale-up of ART gained momentum, facilitated by partnerships with international organizations and donors.^5^ In 2004, Mozambique was among the first countries to benefit support from the United States President’s Emergency Plan for AIDS Relief (PEPFAR), the largest commitment by any nation to address a single disease.^6^ Under the direction of the Government of Mozambique and in collaboration with other partners, PEPFAR has provided resources for prevention, care, and treatment programs, which has also strengthened the country’s capacity to address epidemics and improve health outcomes.^78^ Programmatic innovations, such as the “test and start” strategy (i.e., promptly initiating all newly identified PLHIV on ART) introduced in 2016, and new treatment regimens, such as dolutegravir (DTG)-based regimens introduced in 2019, have improved the reach and effectiveness of ART.

To describe the impact of PEPFAR-supported ART in Mozambique since 2004, we analyzed the number of PLHIV on ART and viral load suppression (VLS) rates as a proxy for understanding treatment effectiveness, and estimated the number of new HIV infections prevented and deaths averted, largely due to the scale-up of ART.

## Methods

We analyzed routine PEPFAR program Monitoring, Evaluation, and Reporting (MER) program data to describe ART scale-up in Mozambique between 2004–2023. Publicly available archival reports from the Mozambique Ministry of Health National HIV Program were also sourced.

In general, PEPFAR defines PLHIV on ART as the number of adults and children who are currently receiving ART in accordance with the nationally approved treatment protocol at the end of the reporting period. The cut off for defining “on ART” has evolved. When ART began in 2004, clients were considered “on ART” if they had an ART pharmacy pick-up within ≤90 days since their last scheduled visit. In October 2018, this changed to ≤28 days since their last scheduled visit. Estimated ART coverage was calculated using the 2024 UNAIDS Spectrum estimates. Data on viral load results were available from 2016 for sites receiving direct PEPFAR support. VLS rates were calculated as PLHIV at PEPFAR-supported sites with a test result indicating VLS (<1,000 viral copies/mL) among those tested in the previous 12 months. PLHIV on ART and VLS were analyzed for children (<15) and for adults (≥15), by sex, and by province. The fiscal years are October through September.

The 2024 UNAIDS Spectrum AIDS Impact Model (AIM) and Goals Age Structured Model (ASM) was used to estimate the number of infections averted, including among HIV-exposed infants, and HIV-related deaths averted by mid-year (July–June) for 2004 to 2023. This model uses national program statistics, survey and surveillance data, and study-derived epidemiologic parameters to calibrate structured models of HIV transmission and produce indicators such as incidence and mortality.^9^ To estimate the number of infections and deaths averted in the absence of PEPFAR, the pattern of infection and deaths that was already occurring before the existence of ART was used to extrapolate the number of deaths and infections which would have occurred in the country without the introduction of ART. This activity was reviewed by CDC, deemed not research, and was conducted consistent with applicable federal law and CDC policy.<u>^1^</u>

## Results

From September 30, 2004, to September 30, 2023, the number of PLHIV on ART in Mozambique increased 5,779 times (from 355 to 2,051,626) (Figure 1), and estimated ART coverage increased from 0.61% to 80.73%. The number of adults living with HIV on ART increased 8,638 times, from 226 to 1,952,272 (85.67% estimated ART coverage) and the number of children living with HIV on ART increased 766 times, from 129 to 98,850 (64.7% estimated ART coverage). In the same time period, the number of women living with HIV on ART increased 3,970 times (from 342 to 1,357,661), while the number of men living with HIV on ART increased 53,382 times (from 13 to 693,965). By September 2023, the number of PLHIV on ART varied among the 11 provinces, ranging from 59,948 (89% estimated ART coverage) in Niassa to 433,016 (96% estimated ART coverage) in Zambezia.

**Figure 1:**
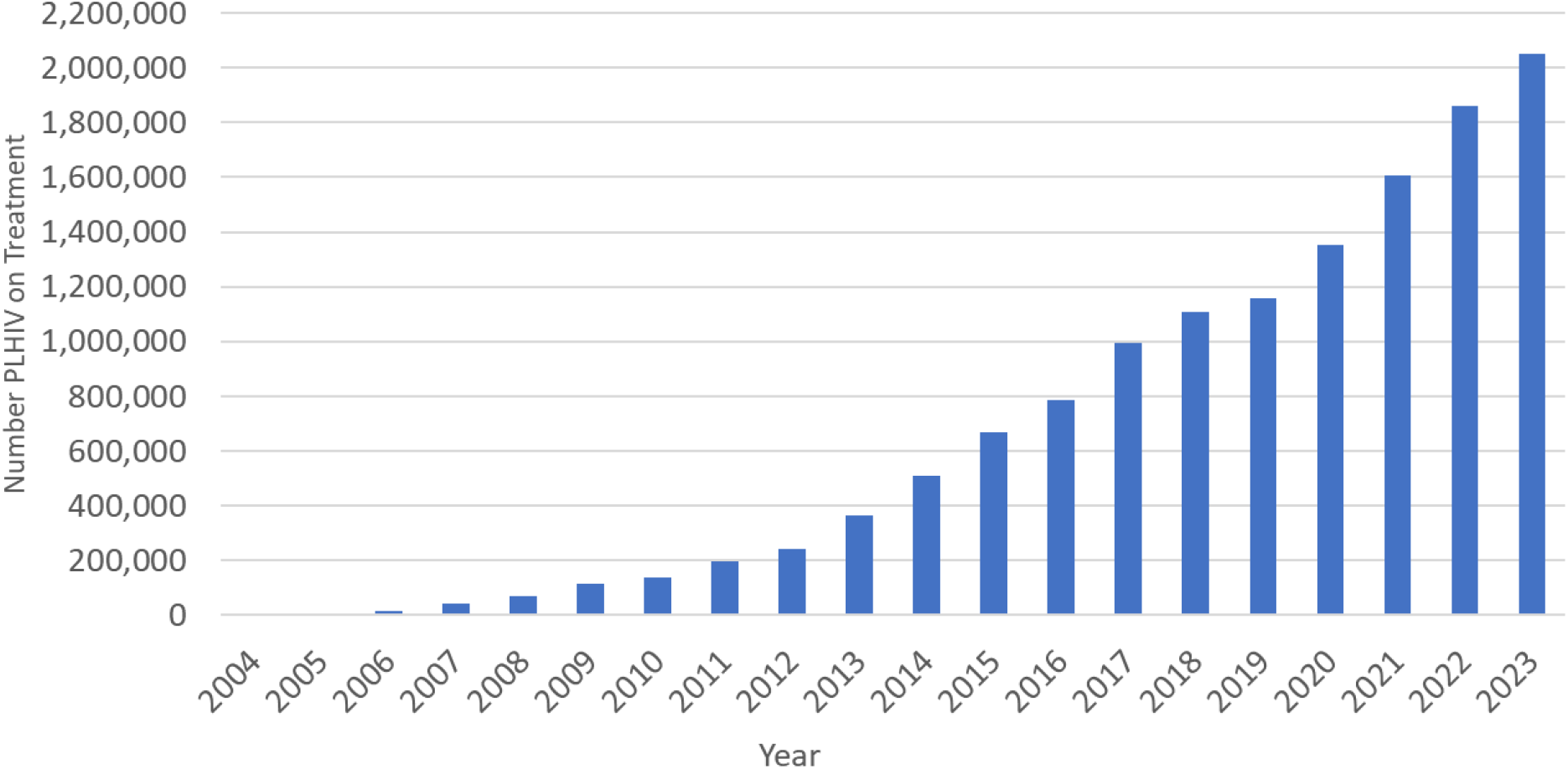
Number of PLHIV on Treatment in Mozambique (2004-2023)

In fiscal year 2016, 174 (23.5%) of 740 PEPFAR-supported sites reporting PLHIV on ART also reported data on viral load, comprising 9,030 (1%) of the 787,612 total PLHIV on ART; in 2023, 636 (100%) of sites reported data on viral load, comprising 1,318,544 (64%) of the 2,051,626 total PLHIV on ART. Between 2016-2023, the number of viral load tests conducted among PLHIV on ART at PEPFAR-supported sites increased 146 times, (from 9,030 to 1,318,544), and the VLS rate increased from 77% (6,954 suppressed of 9,030 tested) to 94% (1,235,626 of 1,318,544) (Figure 2). For clients tested between September 2022 and September 2023, VLS rates were higher among adults (94% [1,178,751 of 1,252,659]) than among children (86% [56,871 of 65,880]), and higher among women (94% [841,115 of 896,120]) than among men (93% [394,511 of 422,424]).

**Figure 2:**
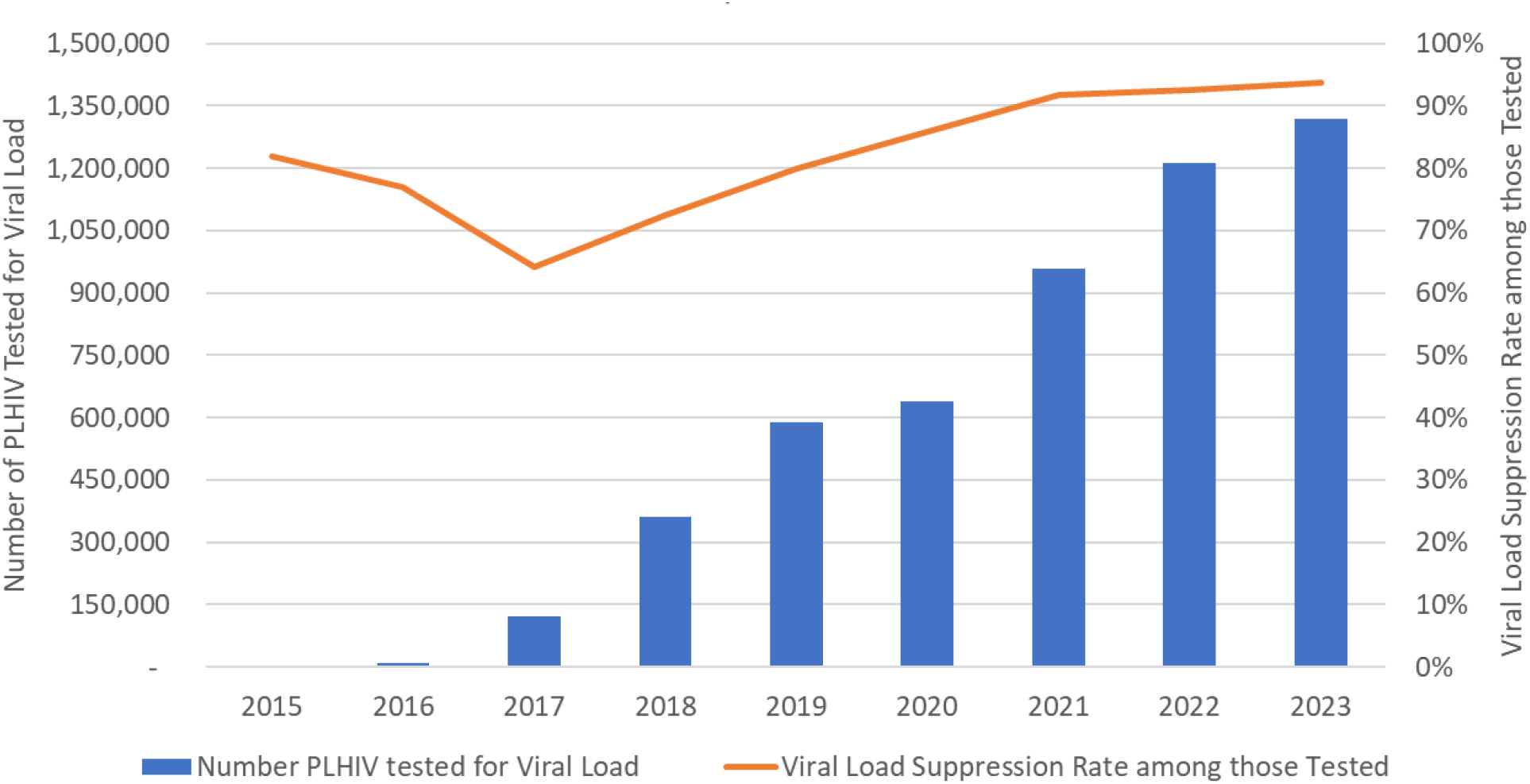
Viral Load Testing Scale-up and Viral Suppression Rates in Mozambique 2015-2023

From Spectrum estimates, in 2004, there were an estimated 1,171,321 PLHIV, with 155,480 new infections; in 2023, there were an estimated 2,438,685 PLHIV, with 81,151 new infections. From 2004 to 2023, an estimated 1,600,000 HIV infections were averted, including 352,256 among HIV-exposed infants. Annually, a median of 60,000 HIV infections were averted, ranging from 1,400 in 2004 to 197,803 in 2022 (Figure 3). During this period, an estimated 1,070,295 total deaths were averted, ranging from 1,514 in 2004 to 95,112 deaths in 2023 (annual median = 53,057).

**Figure 3:**
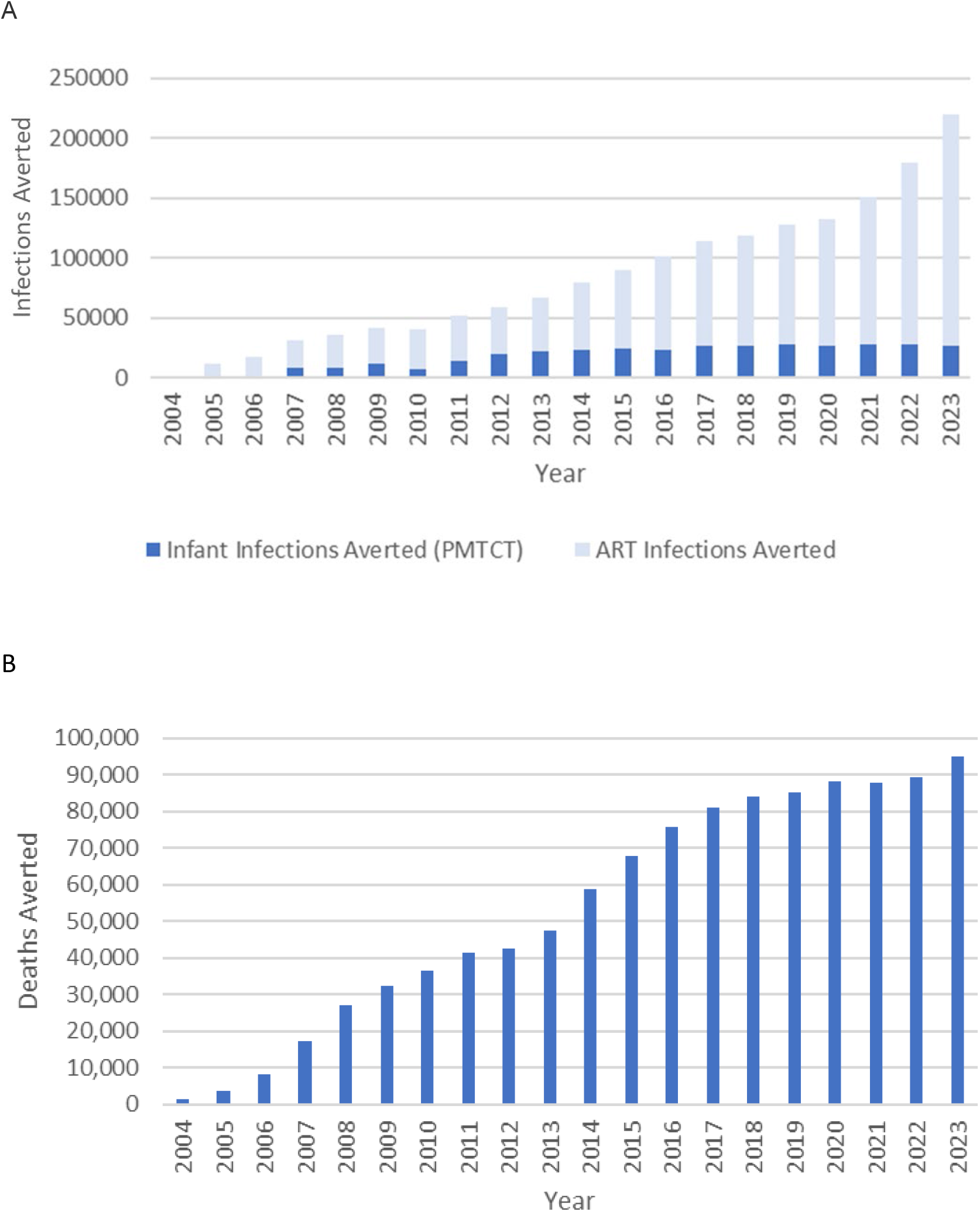
Total of Infections (A) and Deaths (B) Averted, Mozambique 2004-2023

## Discussion

Over the past 20 years, Mozambique has achieved remarkable gains toward ending HIV as public health threat, through the scale-up of life-saving ART, reaching over 2 million PLHIV by September 2023. ART is effective, with 94% of those with a viral load test being suppressed. Additionally, the scale up of ART has averted an estimated 1.6 million HIV infections and over one million deaths. Despite these extraordinary gains, challenges remain, as progress has not been uniformly shared across all ages, sexes, and geographic areas. Children are less likely to be diagnosed and have continuously had worse treatment outcomes as compared to adults, including lower VLS rates, as seen in our analysis.^10,11^

Similar to other sub-Saharan African countries, men in Mozambique have worse HIV outcomes compared to women, including lower VLS rates, as seen in our analysis.^6^ Men are more likely to die of an HIV-related disease than women despite higher incidence and prevalence rates among women.^12^ In part, worse treatment outcomes among men stem from lower uptake of HIV testing services, which leads to diagnosis at later stages of HIV illness, as indicated by higher rates of advanced HIV disease among men. Comparative worse treatment outcomes are particularly pronounced among adolescent and young men.^13^ In addition to poor health-seeking behaviors and attitudes about masculinity, health policies framework in sub-Saharan Africa are largely built to support women of reproductive age and don’t integrate men’s health needs. Therefore services are not seen as friendly to men’s needs.^14^

Finally, there are substantial provincial variations in estimated ART coverage and VLS rates across Mozambique.^9^ Southern provinces consistently outperform both central and Northern provinces, not just in the HIV response but in broader health metrics.^15^ Furthermore, northern Mozambique is experiencing a humanitarian crisis, which has an estimated 542,000 Displaced Population in Cabo Delgado on ART as of January 2024.^16^ The unrest has created an extremely challenging environment to manage chronic diseases, including HIV.^17^ Finally, in recent years, the intensity and frequency of catastrophic climatic events has exacerbated challenges for Mozambique, due to the infrastructure needed to deliver and access quality health services.^18,19^

To address such challenges, Mozambique’s HIV Program is using data to continually improve ART coverage and outcomes and to close equity gaps for all sub-populations and geographic areas affected by HIV. The HIV Program aims to increase access to patient-centered ART delivery options for the majority of ART clients who are stable, including through multi-month drug dispensing of optimized, dolutegravir-based regimens. Patient-centered approaches not only benefit the general PLHIV population, but also allow for tailored services for clients in sub-populations with historic challenges, such as children, youth, men, and key populations.^20^ In addition, continued involvement of communities and civil society to design and monitor the quality of patient-centered programs and other efforts to reduce stigma and discrimination remain vital, as well as those to address social and structural determinants of health, including overall health literacy, infrastructure, and access to facility-based care. While treatment is a key prevention strategy,^21^ concurrent other combination prevention options are important to further reduce the number of people newly infected with HIV who require diagnosis and linkage to lifelong treatment. Finally, health systems strengthening remains both a priority and a challenge, including timely access to viral load testing, trained health care workers, a stable supply chain, and high-quality data, longitudinal data, including on HIV drug-resistance.

These findings have several limitations. First, indicator definitions and systems to report data in Mozambique have changed over time, which might have affected data quality despite continual PEPFAR and national data quality assurance activities. Second, PLHIV can access health services at any site, regardless of residence and therefore, some persons might have been counted more than once. Third, the model estimated averted HIV infections and HIV-related deaths based on ART; however, other services (e.g., voluntary medical male circumcision) and contextual factors beyond ART scale up might have contributed. Fourth, VLS rates were calculated for sites receiving direct PEPFAR support, and overall VLS rates might differ. Finally, it is not possible to quantify the contribution from PEPFAR and other stakeholders (e.g., UNAIDS and the Global Fund) in support of the Government of Mozambique to scale up ART, because investments in infrastructure, leadership, and financing (including commodities) have worked synergistically with PEPFAR investments and programming.

## Conclusion

Between 2004–2023, the scale up of ART reached more than 2 million PLHIV in Mozambique, helping to avert over 1.6 million new infections and over one million HIV-related deaths. Going forward, efforts will focus on covering gaps toward reaching and sustaining HIV epidemic control, including identifying all sub-populations of PLHIV to tailor services and rapidly link them to effective and durable ART.

## Data Availability

All data produced are available from PEPFAR Monitoring, Evaluation, and Reporting program data.

https://data.pepfar.gov/datasets#PDD

## Acknowledgements

The authors would like to acknowledge the contribution of the health care providers in Mozambique who are working daily in the field providing care to patients, as well as PEPFAR implementing partners who have been providing technical assistance to the health facilities. We are also grateful for the support of Margaret Gutierrez-Nkomo and Kwame Asamoa for their critical review to the manuscript.

## Funding Acknowledgement

This manuscript has been supported by the President’s Emergency Plan for AIDS Relief (PEPFAR), through the U.S. Centers for Disease Control and Prevention (CDC), The findings and conclusions in this manuscript are those of the author(s) and do not necessarily represent the official position of the funding agencies.

## Footnotes

1 See e.g., 45 C.F.R. part 46, 21 C.F.R. part 56; 42 U.S.C. §241(d); 5 U.S.C. §552a; 44 U.S.C. §3501 et seq.

